# A mathematical model for multiple COVID-19 waves applied to Kenya

**DOI:** 10.1101/2023.09.01.23294943

**Authors:** Wandera Ogana, Victor Ogesa Juma, Wallace D. Bulimo, Vincent Nandwa Chiteri

**Affiliations:** African Mathematics Millennium Science Initiative, Nairobi, Kenya; Department of Mathematics, University of Nairobi, Nairobi, Kenya; Mathematics Department, University of British Columbia, 1984 Mathematics Road, Vancouver, BC, V6T 1Z2, Canada; Centre for Virus Research and the Department of Epidemiology, Statistics and Informatics, Kenya Medical Research Institute, Nairobi, Kenya

**Keywords:** Mathematical model, COVID-19 pandemic, non-pharmaceutical interventions, delay functions, multiple waves

## Abstract

The COVID-19 pandemic, which began in December 2019, prompted governments to implement non-pharmaceutical interventions (NPIs) to curb its spread. Despite these efforts and the discovery of vaccines and treatments, the disease continued to circulate globally, evolving into multiple waves, largely driven by emerging COVID-19 variants. Mathematical models have been very useful in understanding the dynamics of the pandemic. Mainly, their focus has been limited to individual waves without easy adaptability to multiple waves. In this study, we propose a compartmental model that can accommodate multiple waves, built on three fundamental concepts. Firstly, we consider the collective impact of all factors affecting COVID-19 and express their influence on the transmission rate through piecewise exponential-cum-constant functions of time. Secondly, we introduce techniques to model the fore sections of observed waves, that change infection curves with negative gradients to those with positive gradients, hence, generating new waves. Lastly, we implement a jump mechanism in the susceptible fraction, enabling further adjustments to align the model with observed infection curve. By applying this model to the Kenyan context, we successfully replicate all COVID-19 waves from March 2020 to January 2023. The identified change points align closely with the emergence of dominant COVID-19 variants, affirming their pivotal role in driving the waves. Furthermore, this adaptable approach can be extended to investigate any new COVID-19 variant or any other periodic infectious diseases, including influenza.

## 1 Introduction

COVID-19 is a disease caused by the novel coronavirus SARS-CoV-2 that emerged at the end of December 2019 and has since spread globally. The disease has had an adverse impact on the socioeconomic and health structures of many countries. In Kenya, the virus was first detected on 13^th^ March 2020. Soon after, the Kenyan government implemented non-pharmaceutical interventions (NPIs) to slow the spread of the disease, including the closure of learning institutions, limiting crowding in public transport vehicles and other places to enforce social distancing, and mask-wearing in public areas, etc. Due to low compliance by the public and a rapid rise in infection, the government imposed more stringent measures for instance, banning political and social gatherings, country-wide overnight curfew, suspension of international air travel, closure of bars and clubs, and closure of places of worship, among others. The government additionally imposed COVID-19 regulations, the violation of which constituted a criminal penalty [1]. These measures remained in effect from 8^th^ April 2020 to 8^th^ June 2020. Enforcement of the measures adversely affected the country’s economy and people’s livelihoods. Consequently, the government gradually relaxed some of the mitigation measures during the period 9^th^ June 2020 to 8^th^ August 2020; for instance, places of worship opened with limited numbers of congregants, the lockdown was lifted in parts of Nairobi, Mombasa and Mandera, and there was resumption of international air travel. The daily infections began to go down but suddenly they started to increase in late 2020, fuelled largely by the introduction of a new variant of COVID-19 and partly by the reopening of learning institutions. The rise in infection became so concerning that a lockdown was enforced on five counties, including Nairobi and its environs, in April and May 2021. Luckily, additional mitigation measures became available following the commencement of vaccination in March 2021. The pandemic continued to oscillate in uneven waves of varying amplitudes, as a result of the emergence and spread of new variants of concern.

Following the emergence of COVID-19, future waves were primarily driven by developing variants of concern [2, 3], NPIs [4, 5], vaccines and therapy [6, 7], human behaviour [8, 9], health system status [10] and host sensitivity to the virus and disease [11–14]. Some, if not all, of these factors, should be incorporated in any investigation concerning the dynamics of COVID-19 waves. As a result, COVID-19 has stimulated extensive research by collaborators from many disciplines determined to address the challenges posed by the pandemic. One such challenge involves the use of mathematical modelling to analyse, predict and simulate the dynamics of the pandemic, taking into consideration the myriad drivers of the waves. Modelling of COVID-19 is an active area of research that involves many varied approaches, as can be seen from a recent extensive review [15]. We will concentrate on compartmental models, which are the source of our current contribution. According to these models, the human population is usually divided into five compartments, namely Susceptible (S), Exposed (E), Infected (I), Recovered (R) and Dead (D) [16, 17]. By considering the rate of change of individuals in a compartment, and the contribution of appropriate compartments to this change, we obtain a system of five ordinary differential equations, including parameters that define the rate of flow between adjacent compartments. Models that use all of the compartments are named SEIRD, but those that omit the Dead compartment are called SEIR, and those that omit the Exposed compartment are labelled SIRD. If the Dead compartment is omitted, we could also end up with the classic SIR model, where R here refers to removed, namely those who have recovered or are dead. In order to incorporate the effects of various drivers of COVID-19, additional compartments may be created and appropriate interactions defined. This leads to more complicated systems with more ordinary differential equations and increased numbers of parameters, thus increasing the level of difficulty of solving the equations [18–22]. Compartmental models can also be formulated for more complex problems and the findings can serve as a guide to policymakers, as illustrated by the models for China [23], United Kingdom [5], Ukraine [24] and USA [25]. Models have also been developed that address limited issues in Kenya, for instance, [26–29].

The classical SEIRD model and its derivatives are designed to yield results in a single wave, since the computed infection curve is smooth and has only one peak. Observed infection curves on the other hand are not smooth as they depict many spikes and sub-epidemics. The objective of incorporating the COVID-19 drivers in compartmental models is to try and replicate the spikes and sub-epidemics, as much as possible. Some models have been modified to achieve this replication and, in addition, attempt to forecast multiple waves, as shown in the following examples. Kaxiras and Neofotistos [30] used the SIR model to investigate the effects of social distancing, with a focus on identifying features that can emulate real data. They used a microscopic model in which an infected individual can only infect other individuals within a range in the neighbourhood. The model attempts to explain spikes in a wave but there is no evidence that it can forecast another wave. Perakis et. al. [31] apply the discrete version of the SEIRD model and identify a time, called the change point, that marks the end of one wave and the beginning of the next. They postulate that the recovery and infection rates have jump values at the change point. Using infection data, they apply martingales to identify the change point and hence evaluate the associated jump values of the parameters. The method describes well the spikes in one wave but it is less successful in generating another wave. The main basis of the approach by Ghosh and Ghosh [32] is that the susceptible fraction increases now and again by multiple re-infection of people who have recovered. To achieve this, they add a delay term of the infection, multiplied by a parameter regarded as the rate of re-susceptibility, to the equation involving the rate of Susceptible. The same term is subtracted from the equation involving the rate of change of the Removed. To avoid obtaining a purely periodic solution, they assign suitable values to the transmission rate, depending on the time relative to the delay constant, while holding the removal rate constant. The results provide a good match for the data from India during the selected waves. Leonov et. al. [33] use the SEI model, with the parameters assumed to be piecewise constant, rather than constant as in the classical case. They add an arbitrary function of time to the equation involving the rate of change of infection. This additional term can be considered as a source of infections associated with all other miscellaneous sources. The final solution is obtained from the inverse problem. The method replicates spikes well but it leads to large errors when the gradients of the infection are large, as would be the case involving a new wave.

To replicate observed infection curves, some compartmental models, as pointed out above, include various drivers of COVID-19 dynamics in additional compartments. These drivers can consist of a mitigation force, namely, a force that reduces the transmission rate of the disease, like the application of vaccines; or it can consist of a relaxation force, namely a force that increases the transmission rate of the disease, like increased crowding at a rally or stadium. Since there are so many drivers, it would be unrealistic to attempt to account for them all [31]. There exist some compartmental models, however, that consider the total mitigation and postulate that its effect on the transmission rate can be represented by a piecewise continuous function involving exponential, logistic, linear or constant functions [30, 34–37]. This concept was extended by Ogana et. al.[26] to include relaxation so that the effect of mitigation and relaxation forces on the transmission rate could result in a piecewise exponential-cum-constant function, where the exponential function decreases for mitigation, and increases for relaxation. They applied this method to the SIRD system to compute the first COVID-19 wave in Kenya. The method is simple and flexible and can easily be applied to examine different scenarios, pending more rigorous investigation on the effect of specific drivers of the pandemic.

The current paper uses the method in [26] together with entirely new concepts, as described hereunder. We noted that the solution of the SIRD system, in the absence of any subsequent interventions, has a computed infection curve which dissipates with time. Furthermore, a new wave is formed when the observed infection curve diverges from the dissipating computed infection curve. We chose the”change point”, namely, the boundary between successive waves, according to Perakis et. al [31], among others, as the time at which this divergence commences. We were able to establish that at a point on the computed wave, where infection decreases, it is possible to computationally generate a new wave by application of an appropriate relaxation force in the neighbourhood of the change point. The relaxation strength can be adjusted so that the fore section of the generated wave closely matches the shape of the observed infection curve. Some observed infection waves have very low infection fractions, near the change point, with curves that are approximately horizontal. Furthermore, the susceptible fraction for such waves is approximately constant thus making it possible to approximate the fore section of the infection curve by an exponential function. Once the fore section of the current computed wave is generated, we choose a point on it, that also lies on, or close to, the observed infection curve. We then replace the disease variables at the chosen point on the fore section with the variables at a point in a previous wave, where the infection equals that at the chosen point on the fore section. Since the two points have the same infection value and the same direction of infection, there must be a similarity in the dynamics of the disease in the neighbourhoods of the two points. This procedure introduces a jump in the susceptible fraction and perpetuates the growth of a wave in the right direction. Mitigation or relaxation is then undertaken to yield the complete computed infection curve for the wave. Using these findings, we have computed waves that replicate the observed COVID-19 waves in Kenya to date. In addition, we determine the magnitudes of the major mitigation and relaxation forces associated with changes in the waves.

We present the paper according to the following outline. In Section 2.1, we describe the SIRD model. Section 2.2 contains the derivation of the equations that describe the effects of the intervention on the transmission rate. Section 2.3 has a review of the results for the first wave as obtained in a previous publication. In section 2.4 we present the detailed models of the second to fifth waves, starting with the generation of a wave through the application of a sufficiently large relaxation. In Section 2.5 we present the detailed models of the sixth and seventh waves, starting with the generation of a wave by assuming exponential infection. Section 3.1 contains results and a discussion of the modelled 1st wave compared with observation. Section 3.2 contains results and a discussion of the modelled 2nd to 5th waves compared with observation. Section 3.3 contains results and a discussion of the modelled 6th and 7th waves compared with observation. Finally, we give a few concluding remarks and recommendations in Section 4.

## 2 METHODS

In this section, we first present the equations for the SIRD model and then derive the equations that describe the effects of the intervention on the disease transmission rate. We present computational results of the effect of applying large relaxation forces at a point where infection decreases, particularly in the neighbourhood of transition from one wave to another. The results form the basis of generating the first five waves of COVID-19 in Kenya. Finally, we derive the equations for the generation of the 6^th^ and 7^th^ waves.

### 2.1 Baseline Dynamics by SIRD Model

The dynamics of COVID-19 can be investigated by a variety of methods including compartmental models in which, at the time, *t*, the population is divided into five basic classes, namely: Susceptible, Exposed, Infected, Recovered and Dead, denoted by, *S*(*t*), *E*(*t*), *I*(*t*), *R*(*t*) and *D*(*t*), respectively. Depending on the phenomena being investigated, some compartments may be excluded or new compartments may be added. In this paper, we will apply the SIRD model in which the Exposed (*E*) component is omitted, as shown in Figure 1.

**Figure 1:**
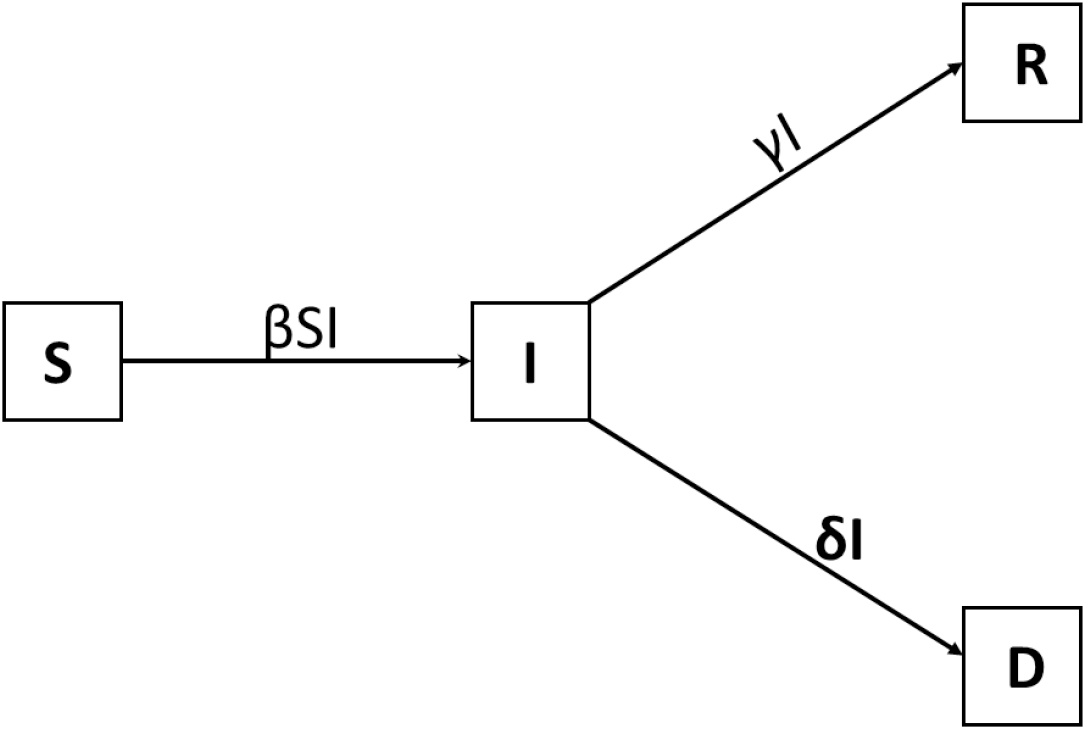
Compartmental SIRD model.

We assume that the total population, *N*, is constant over time. For simplicity, the variables are already normalised on division by *N* such that

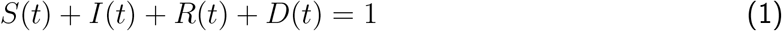

*S*(*t*), *I*(*t*), *R*(*t*) and *D*(*t*) now represent the fractions or proportions of the Susceptible, Infected, Recovered and Dead in the population, at any given time *t*. The governing differential equations are given as follows (see for example, [16, 26]).

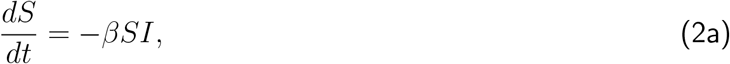

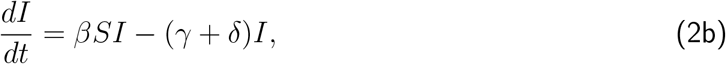

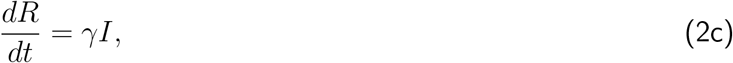

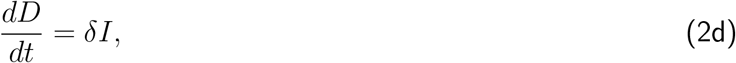

subject to the initial conditions:

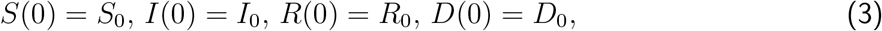

where *S*_0_, *I*_0_, *R*_0_ and *D*_0_, are the initial fractions of the Susceptible, Infected, Recovered and Dead, respectively.

The system of equations (2) contains the parameters: *β*, the disease transmission rate; *γ*, the recovery rate; and *δ*, the death rate. Solution of System (2) together with initial conditions (3) involves determining the parameters *β, γ*, and *δ* that lead to the minimization of some error norm. COVID-19 was first detected in Kenya on 13^th^ March 2020 and it spread till 8^th^ April 2020, before any measures were undertaken to control its spread. This period serves as the reference timeframe during which the disease spread without any intervention, and hence we refer to it as the baseline period. With the death rate, *δ* = 0.015, approximated from the Case Fatality Rate (CFR), minimization yielded the following values for the other 2 parameters during the baseline period [26].

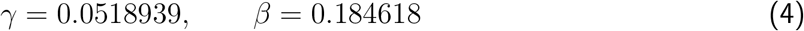

From Equation (4) we obtain the reproduction number, *R*_0_ = 2.76 and the recovery days, 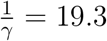 If *γ* and *β* in Equation (4) are substituted into Equation (2) one can determine the trajectory of the infected fraction, among the other variables. The model we develop in the current paper uses the values in Equation (4) and assumes that interventions affect only the transmission rate, *β*, while the recovery and the death rates remain constant.

### 2.2 Effect of Interventions on the Transmission Rate

There exist two types of forces in epidemic interventions: mitigation forces that reduce the rate and extent of infection and relaxation forces that increase the rate and extent. Although mitigation forces are generally perceived as non-pharmaceutical interventions (NPIs), arising from implementation of government policy, we will include among them medical treatment and vaccination, as these actions help to reduce disease spread. Relaxation forces are generally perceived as the lifting of mitigation measures in order to limit the impact of the disease on society. We will include among them non-compliance with mitigation measures and the emergence of new variants of COVID-19, since both of these dramatically increase the spread of the disease. As noted earlier, there are models that introduce more compartments in order to incorporate mitigation and relaxation forces. Our approach is to consider the totality of mitigation or relaxation forces without isolating individual components.

In this paper, we formulate an intervention model which leads to piecewise exponential-cum-constant functions for the transmission rate, as a result of the effects of interventions. It takes into account the fact that intervention not only leads to a reduction of the transmission rate, through mitigation, but also can lead to a surge in the transmission rate, through relaxation. Let the daily events be at the time nodes denoted *t*_0_, *t*_1_, *t*_2_, *· · ·*. Suppose any intervention (mitigation or relaxation), is initiated at the time node *t*_*k*_ then there will be a difference in the transmission rate before and after *t*_*k*_. Let *β*_*b*_(*t*) be the incoming transmission rate at the time *t*_*k*_. We assume that for any time *t* > t_*k*_, the rate of change of the transmission rate, as a result of intervention, is proportional to the transmission rate at that time. This gives the transmission rate as an exponential function of time. The main objective is to gradually change the incoming transmission rate, *β*_*b*_(*t*), by a fraction *c* so that the transmission rate at a future time, say *t*_*k*+*m*_, where *m >* 0, reaches an optimum value (1 *− c*)*β*_*b*_(*t*_*k*_). It was shown that this yields [26]:

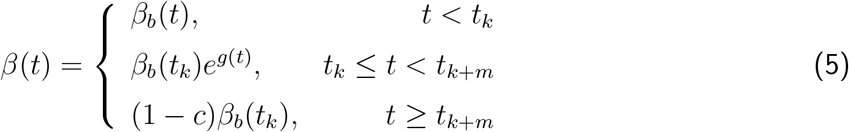

Where

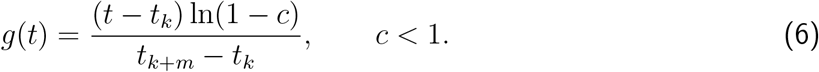

In solving Equation (2), we need to take into consideration the value of the transmission rate, *β*(*t*), according to Equations (5) and (6), depending on whether the time, *t*, is; (i) before an intervention takes place, (ii) after the intervention but before the optimum value is reached ; or (iii) after the optimum value has been achieved. The parameter *m* can be considered as the duration, in days, for the transmission to achieve the optimum value as a result of the intervention.

From the last part in Equation (5) we note that when 0 < *c* < 1, then *β*(*t*_*k*+*m*_) < β>(*t*_*k*_); this corresponds to the intervention being a mitigation, since it yields a smaller future transmission rate which represents a reduction by a fraction *c* of the incoming transmission rate. We call the quantity 100*c* the”percent mitigation”. On the other hand when *c <* 0 then *β*(*t*_*k*+*m*_) > *β*(*t*_*k*_); this corresponds to the intervention being a relaxation since it yields a larger future transmission rate which represents an increase by a fraction |*c*| of the incoming transmission rate. We call 100 *×* |*c*| the”percent relaxation”. If *c* = 0 then *β*(*t*) = *β*_*b*_(*t*_*k*_) for *t* > t_*k*_. This implies that no intervention has taken place at *t*_*k*_, since the incoming transmission rate remains unchanged after the supposed intervention. If this transmission rate is used in solving Equation (2), the resulting infection curve, when *t* > t_*k*_, is called the non-intervention curve. We shall see later that the non-intervention curve plays a significant role in identifying the type of intervention appropriate in modelling COVID-19 waves. Previous researchers restricted *c* to the interval (0, 1); hence they covered only mitigation forces. Through Equation (6), we extend *c* to negative values to account for spikes in the dynamics that may occur as a consequence of relaxation forces. This opens the way for computationally generating new COVID-19 waves.

### 2.3 First wave

Ogana et. al. [26] developed the model of the first wave in three distinct phases: baseline, mitigation and relaxation. We present a summary here because the methods and the findings are fundamental to developing the models of subsequent waves. The baseline phase is covered in Section 2.1. The mitigation phase commenced on 9^th^ April 2020, following mitigation measures announced the day before. In modelling this phase, the best agreement with data was obtained by using, *β*_*b*_(*t*) = 0.184618, *m* = 15, and *c* = −0.41, (41% mitigation) in Equation (5). The relaxation phase commenced on 9^th^ June 2020, following the lifting of some mitigation measures the day before and continued until early August. In modelling this phase, the best agreement with data was obtained by using *β*_*b*_(*t*) = 0.108925, *m* = 15 and *c* = −0.24, (24% relaxation) in Equation (5). After solution of Equation (2), consolidation of results from the three phases leads to Figure 2 in which the modelled percent infection during the first wave is compared with data. Figure 2. also shows the beginning of the observed second wave.

**Figure 2:**
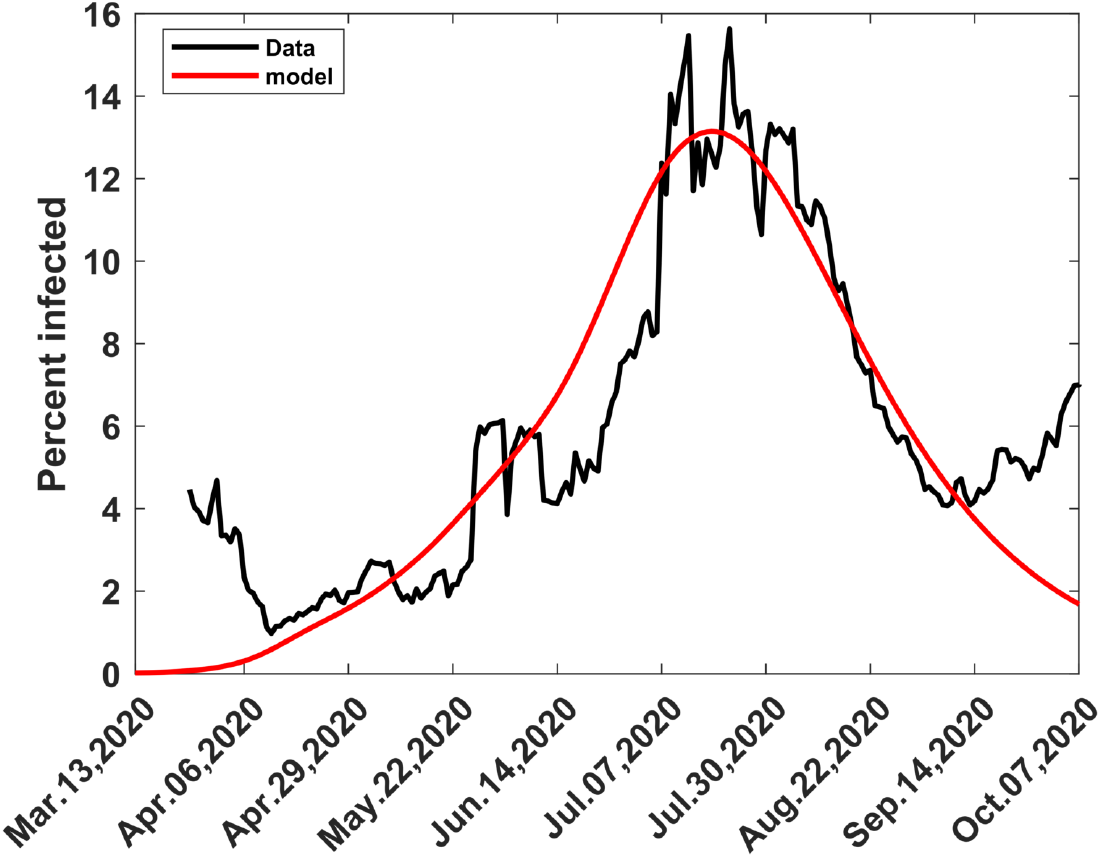
Computed and observed first wave of COVID-19 in Kenya. *T*_*W*_ is the end of the first wave.

### 2.4 Second to Fifth waves

Modelling of the second to fifth waves is different from modelling the first wave, although they have in common the construction of infection scenarios arising from mitigation or relaxation processes. The principle behind the modelling of the second to fifth waves is the fact that, under suitable conditions, it is possible to generate a wave by applying a large enough relaxation force, when infection is decreasing. The generated wave requires some adjustment, however, to make it replicate the observed wave. We illustrate the process by considering what happens between the first and second waves, with the understanding that equivalent techniques can be employed between any two adjacent waves.

#### 2.4.1 Relaxation at Varying Percentages When Infection Decreases

From Figure 2 it can be seen that the computed first wave dissipates with time; it represents the path the infection would take in the absence of any further interventions, including the formation of the second wave. We note that the second wave is formed when its trajectory diverges from the trajectory of the computed first wave in early September 2020. We estimated that the divergence started around 3^rd^ September 2020; hence this date became the change point between the first and second waves, namely, the time when the first wave ended and the second started. In reality the change is more gradual and takes place over a longer period. We carried out computational experimentation by applying relaxation forces, under several scenarios, with the incoming transmission rate kept at *β*_*b*_(*t*) = 0.135067, as established in [26]. The results are in Figure 3, which shows the effect of varying relaxation forces and the application of delay function from one wave to another. For the first scenario, after trying different values of *m*, we chose *m* = 30 and applied varying values of the relaxation ratio, *c*, in Equation (5).

**Figure 3:**
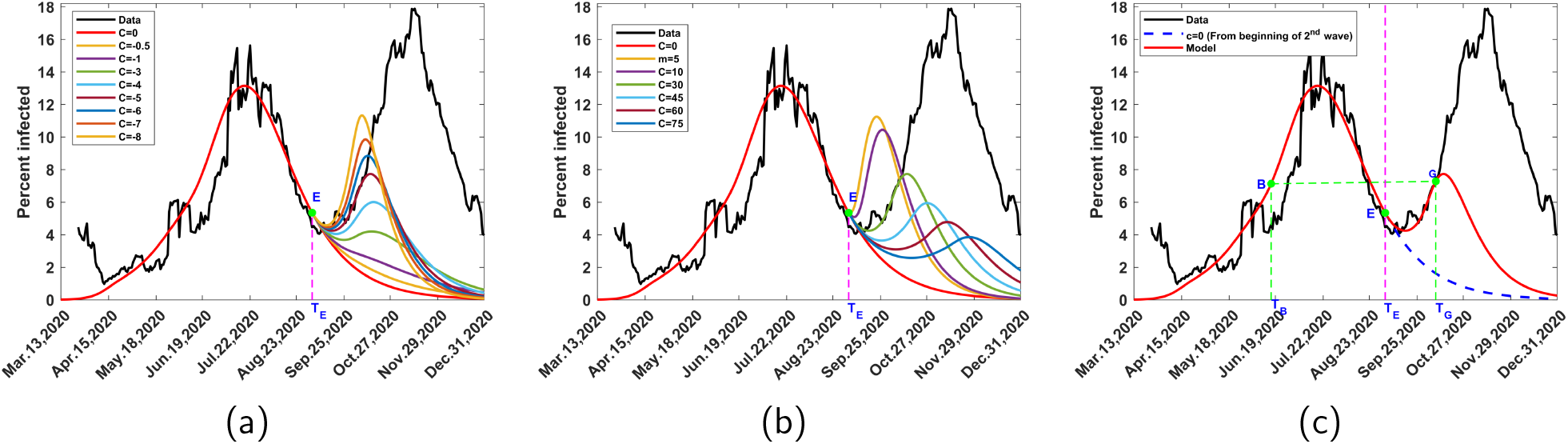
Effects of the applications of relaxations and the application of delay function. E is the Change point from one wave to another, while *T*_*E*_ is the time at E. (a) Relaxations of varying magnitudes when the infection is decreasing; (b) Relaxations at the same magnitude but with varying optimal duration of change of the transmission rate; (c) Choice of points for application of delay function at the current and previous waves. G is the Point of application of delay function condition, while *T*_*G*_ is the corresponding time point; B is Point in previous wave where infection at B equals infection at G and *T*_*B*_ is the time at B.

Subsequent solution of Equation (2), for time after 3^rd^ September 2020, yielded Figure 3a. At low relaxation percentages, there does not appear to be any significant effect, but as the percentages increase, ripples begin to form and eventually turn into sharp waves with large crests. The second scenario concerned the case when the waves are generated at the same relaxation force, namely at a given value of c, but have different values of m, as shown in Figure 3b. As m increases the base of the wave becomes broader while the crests of the waves become smaller and they move downwards. These phenomena can be replicated at suitable points in the subsequent waves, with equivalent results to Figures 3a and 3b. Hence, they form the foundation of our modelling the second to fifth waves as outlined in the next three subsections. Every wave will be divided into the fore and back portions, each having different mechanisms of development.

#### 2.4.2 Fore portion of the wave by relaxation

The computed wave starts with a fore portion generated by application of a sufficiently large relaxation force. The prime candidate for such a force is a new COVID-19 variant. The force could be enhanced by widespread violation of mitigation measures. Let E be the change point between the preceding wave (Wave 1) and the current wave (Wave 2), with the associated time *T*_*E*_ and infection *I*_*E*_ (Fig. 3c). From Figure 3a we have seen that it is possible to generate a series of waves by applying forces of varying relaxation percentages at the point E. To model the fore portion of the current wave (Wave 2), we proceed according to the following steps.

i. Choose the initial change point, *T*_*E*_, as the time when the trajectory of the observed current wave (Wave 2) begins to diverge from the modelled tail of the preceding wave (Wave 1), which dissipates with time, as pointed out in Section 2.4.1.
ii. Choose the default value *m* = 15 and select a coarse grid of negative values of *c*, preferably from the interval [*−*10, 0].
iii. Determine the values at *T*_*E*_ of the transmission rate, *β* and the disease variables *S, I, R* and *D*. Solve Equation (2) for *t > T*_*E*_ while applying Equation (5), with the incoming transmission rate.
iv. Adjust *T*_*E*_ and *m*, if necessary, and repeat step iii., with a finer grid of *c*, till an infection curve is obtained that fits the data well, from the scenario of available curves.

#### 2.4.3 Application of delay function

As Figure 3a shows, the curve obtained in step iv of Section 2.4.2 will reach a maximum and dissipate at large times. To avoid this fate and force the model to follow the data upwards, we pick a point on this curve, which lies on, or closest to the data and is not near the maximum point. Let this point be G, with associated time *T*_*G*_ and infection *I*_*G*_ (Figure 3c). From the preceding wave or any other suitable previous wave, identify a point *B*, on the left side of the wave, with associated time *T*_*B*_ and infection *I*_*B*_. We choose *B* such that *I*_*B*_ = *I*_*G*_, that is, the infection at time *T*_*B*_ equals that at time *T*_*G*_ (Figure 3c). Since the infections at the two points are equal, we assume the similarity of dynamics at the two points and hence require that the rest of the time-dependent variables should also be equal at the two points. We thus impose the condition

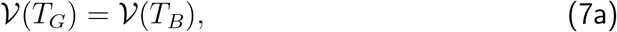

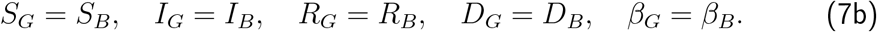

where the subscripts denote the values of the quantities at the corresponding points, as shown in Figure 3c. Equation (7) is known as the delay function condition, since values at the current time, *T*_*G*_, are assigned values at a preceding or previous time, *T*_*B*_. The arc *E −G* is what we refer to as the fore portion of the computed wave. Our objective is to make this arc match the observed infection curve as much as possible, by careful choice of the quantities *T*_*E*_, *m, c, T*_*G*_ and *I*_*G*_. If the whole of the preceding wave falls below or above G, then it is not possible to obtain the point *I*_*B*_ in the preceding wave, such that *I*_*G*_ = *I*_*B*_, hence we seek for *I*_*B*_ from a previous wave.

#### 2.4.4 Back portion of the wave

The back portion of the computed wave begins at the end of the fore portion, namely at *T*_*G*_ and proceeds for *t > T*_*G*_. The fore portion of the wave arises mainly from the effects of the resultant relaxation force due to new COVID-19 variants together with enhanced non-compliance to mitigation measures. The back portion, on the other hand, is influenced by resultant relaxation and mitigation forces as time progresses, mainly due to interventions and the continued effects of the variants. Given the trajectory of the observed infection, it is important to find out whether relaxation or mitigation should be applied at a point of intervention. Suppose the intervention occurs at time *T*_*V N*_ and let the incoming transmission rate of the model be *β*_*b*_. We generate an infection curve from *T*_*V N*_ by using *β*_*b*_ and then compare the trajectory with data in order to determine whether the intervention should be a relaxation or mitigation. This is done by using the following steps.

1. Let the incoming transmission rate at *T*_*V N*_ be *β*_*b*_. Choose the default value m = 15.
2. Choose values of *S, I, R* and *D* at *T*_*V N*_ as initial values. Solve Equation (2) for *t > T*_*V N*_, while adjusting the transmission rate according to Equation (5), with *c* = 0, in order to generate the non-intervention curve (see Section 2.2).
3. Compare the trajectory of the non-intervention curve with data and one of the following two cases will arise:

**Case 1: After** *T*_*V N*_ **the data is predominantly above the non-intervention curve**. This means that the transmission rate for the data is larger than the transmission rate of the model, namely *β*_*b*_. To obtain a model whose transmission rate is close to that of the data, we apply relaxation by choosing negative values of c, according to Equation (5), and use the new transmission rate in solving Equation (2) for *t > T*_*V N*_. By varying *c*, and adjusting *m*, if necessary, we can determine a value of the transmission rate that is close enough to that of the data so as to yield a model infection curve that closely fits the data. For clarity of computation, we let *T*_*V N*_ = *T*_*RX*_ to indicate that the intervention is a relaxation. Furthermore, the relaxation stays in effect until another intervention is encountered.

**Case 2: After** *T*_*V N*_, **the data is predominantly below the non-intervention curve**. This means that the transmission rate for the data is smaller than the transmission rate of the model, namely *β*_*b*_. To obtain a model whose transmission rate is close to that of the data, we apply mitigation by choosing positive values of c, according to Equation (5), and use the new transmission rate in solving Equation (2) for *t > T*_*V N*_. By varying c, and adjusting m, if necessary, we can determine a value of the transmission rate that is close enough to that for the data so as to yield a model infection curve that closely fits the data. For clarity of computation, we let *T*_*V N*_ = *T*_*MT*_ indicate that the intervention is a mitigation. Furthermore, the mitigation stays in effect until another intervention is encountered.

In modelling of the back portion, we may use relaxation, mitigation and delay function, as convenient, to align the model with the data, in the event that the model exhibits departure from the expected trend.

### 2.5 Sixth and Seventh waves

The methods used to generate the fore portions of the 2^nd^ to 5^th^ waves were applied to the 6^th^ and 7^th^ waves but they did not succeed no matter how large a relaxation force was used. We noticed a difference in the formation of the two sets of waves. The 2^nd^ to 5^th^ waves start when a decreasing infection diverges to the right, forms a concave shape, with a base, then increases, as in Figure 2 for the observed 2^nd^ wave. The 6^th^ and 7^th^ waves, on the other hand, emerge from an almost horizontal direction and gradually increase before rising sharply rise, as in Figure 4 on the formation of the 6^th^ wave.

**Figure 4:**
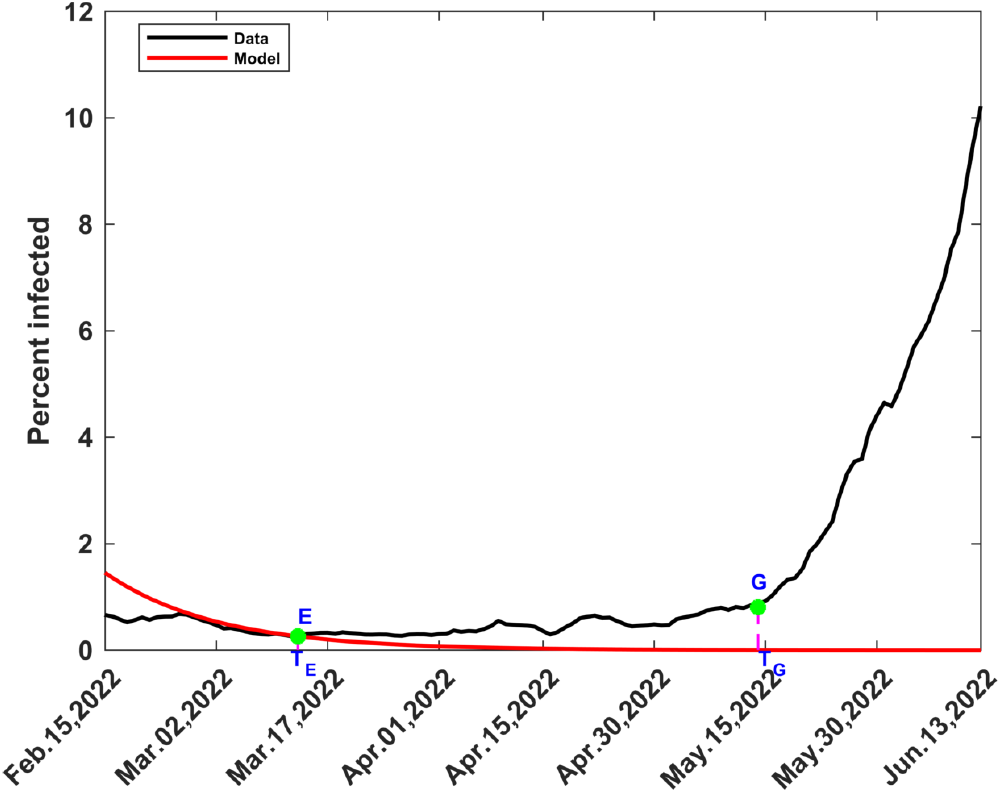
Development of the 6^th^ wave. *E* is the point from 5^th^ to 6^th^ wave and *T*_*E*_ the corresponding time. The black curve represents the data corresponding to the 6^th^ wave, while the red curve corresponds to the computed tail of the 5^th^ wave.

#### 2.5.1 Fore portion of the wave by exponential growth

The fore portions of the 6^th^ and 7^th^ waves can be modelled by exponential infection. The initial change point, *T*_*E*_, between the current and preceding waves, is determined as before by noting when the trajectory of the current observed wave diverges from the modelled tail of the preceding wave, as noted in Figure 4 for the transition from the 5^th^ to the 6^th^ wave. We observed the following properties of the variables *I* and *S* at the beginning of the formation of the 6^th^ and 7^th^ waves:

1. Values of *I*(*t*) were quite small, *<* 1%, in the neighbourhood of the change point, and increased slowly away from the change point.
2. *S*(*t*) was approximately constant for many days after the change point, till the point say *G* in Figure 4, close to where the sharp rise in infection commences.

For our model, the arc *E −G* forms the fore portion of the wave. Using the 2nd property above, we assume that *S*(*t*) is approximated by the mean value,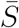, from *T*_*E*_ to *T*_*G*_. Equation (2b) can then be written

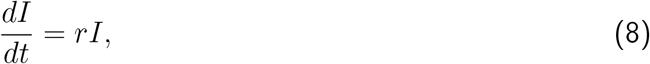

Where

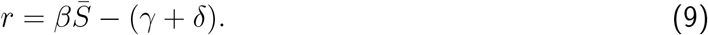

Equation (9) has the solution

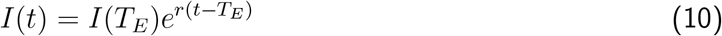

For the infection to grow we must have *r >* 0, implying that we must choose a transmission rate *β* such that,

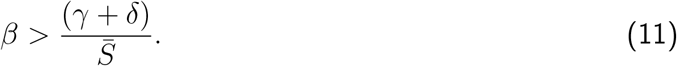

Although Equation (11) gives a wide range of choice for *β*, the value must be selected such that the computed infections from Equation (10) agree with data as well as possible. This is readily done by trying different values of *β* and comparing the exponential curve with data till a suitable value of *β* is reached. We denote such a value *β*_*XP*_, to indicate that it is the transmission rate associated with exponential infection. Using Equation (10), we compute the infected fraction by,

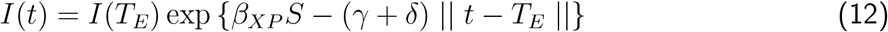

Exponential growth cannot be allowed to proceed indefinitely. Just like in the 2^nd^ to 5^th^ waves, we terminate the fore portions of the 6th and 7^th^ waves by enforcing the delay function condition at the point G, as discussed in Section 2.4.3. Hence Equation (12) is used to determine the infection for *t* such that *T*_*E*_ *≤t ≤T*_*G*_. Within this time interval, 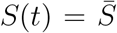, a constant, while *R*(*t*) and *D*(*t*) are estimated from

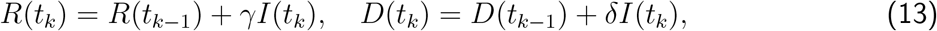

through integration of Equations (2c) and (2d), respectively.

#### 2.5.2 Back portion of the wave

The back portion of the wave starts at *T*_*G*_ and its modelling proceeds as described in Section 2.4.4, with the following points to be noted:

- The first intervention is a relaxation at *T*_*G*_. It produces a model, which replicates the left half of the wave.
- The second intervention is a mitigation, near the apex of the wave, and it produces a model which replicates the right half of the wave.

## 3 RESULTS

Presentation of results here will be done one wave at a time; thereafter all the waves will be consolidated into one time series. COVID-19 data was obtained from the Ministry of Health, Kenya [38] and Worldometer [39]. We obtained information on SARS-CoV-2 lineages and variants from Gathii et. al [3] and Nasimiyu et. al. [40]. After the first usage, we will refer to lineages and variants without attaching SARS-CoV-2 every time.

### 3.1 First Wave

The first wave was modelled by Ogana et. al. [26] and a summary of the results is given in Section 2.3, with the complete wave shown in Figure 2. It was driven largely by the global parental SARS-CoV-2 lineage B.1 that lasted from March 2020 to September 2020 [3, 40]. The fluctuations in the wave were, however, partly influenced by the mitigation measures imposed during 8th April 2020 to 8th June 2020 and the lifting of some of these measures from 8th June 2020. These actions led to unique mitigation and relaxation dynamics different from what would have happened if the disease had been allowed to spread without any intervention [26].

### 3.2 Second to Fifth Waves

We present the results one wave at a time. The procedures are almost identical; the differences occur in the dates when major events and decisions take place, and hence the attendant output.

#### 3.2.1 Second Wave

The methods in Section 2.4.2, led to = 0.13507, *m* = 30, *c* = *−*4 (400% relaxation) and *T*_*E*_ = 03*−*Sep*−*20. From Section 2.4.3, we identified *T*_*G*_ = 08*−*Oct*−*20 and *I*_*G*_ = 0.07394. Comparison with previous waves led to *T*_*B*_ = 17*−*Jun*−*20 and *I*_*B*_ = 0.07394. Equation (7) yielded the values in column 3, Section A2 of Table A1. The fore portion of the wave is the arc *E − G*. The methods in Section 2.4.4, applied at *T*_*V N*_ = *T*_*G*_ = 08 −Oct −20 with *β*_*b*_ = 0.67533, led to the non-intervention curve (blue dashed curve), predominantly below data as shown in Figure 5. Hence we let *T*_*RX*_ = *T*_*V N*_ and undertook final relaxation for *t > T*_*RX*_ using *c* = −0.26 (26% relaxation) and *m* = 15. This gave the back portion of the wave which, combined with the fore portion, yielded the complete 2^nd^ wave shown in Figure 5.

**Figure 5:**
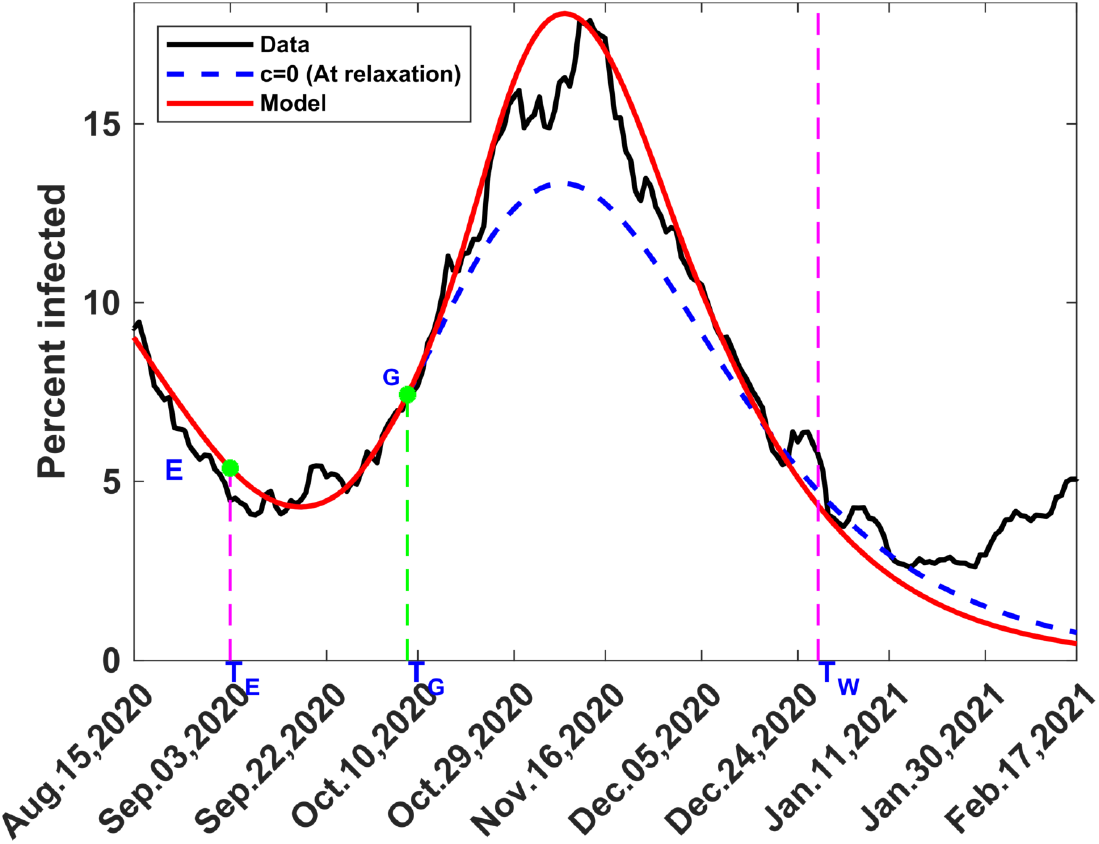
Observed and computed 2^nd^ Wave of COVID-19 in Kenya. The blue curve represents the no-intervention curve, black represents the data while the red is the model. *E* is the change point from first to the second wave, with the corresponding time *T*_*E*_; *G* is the point of application of delay function condition and *T*_*G*_ the corresponding time; *T*_*W*_ shows the end of second wave.

#### 3.2.2 Third Wave

The methods in Section 2.4.2, led to *β*_*b*_ = 0.17018, *m* = 45, *c* = *−*9 (900% relaxation) and *T*_*E*_ = 28*−* Dec*−*20. From Section 2.4.3, we identified *T*_*G*_ = 10*−*Feb*−*21 and *I*_*G*_ = 0.04099. Comparison with previous waves led to *T*_*B*_ = 26 −May −20 and *I*_*B*_ = 0.04099. Equation (7) yielded the values in column 4, Section A2 of Table A1. The fore portion of the wave is the arc *E − G*. Enforcement of relaxation at *T*_*G*_ resulted in a model to the left of data. To align the model with data, we carried out computation for *t > t*_*G*_, with *c* = 0, and chose *T*_*V N*_ = 21*−*Feb*−*21 with *β*_*b*_ = 0.10892. The methods in Section 2.4.4, led to the non-intervention curve (blue dashed curve), predominantly below data as shown in Figure 6. Hence we let *T*_*RX*_ = *T*_*V N*_ and undertook final relaxation for *t > T*_*RX*_ using *c* = − 0.5 (50% relaxation) and *m* = 10. This gave the back portion of the wave which, combined with the fore portion, yielded the complete 3^rd^ wave shown in Figure 6.

**Figure 6:**
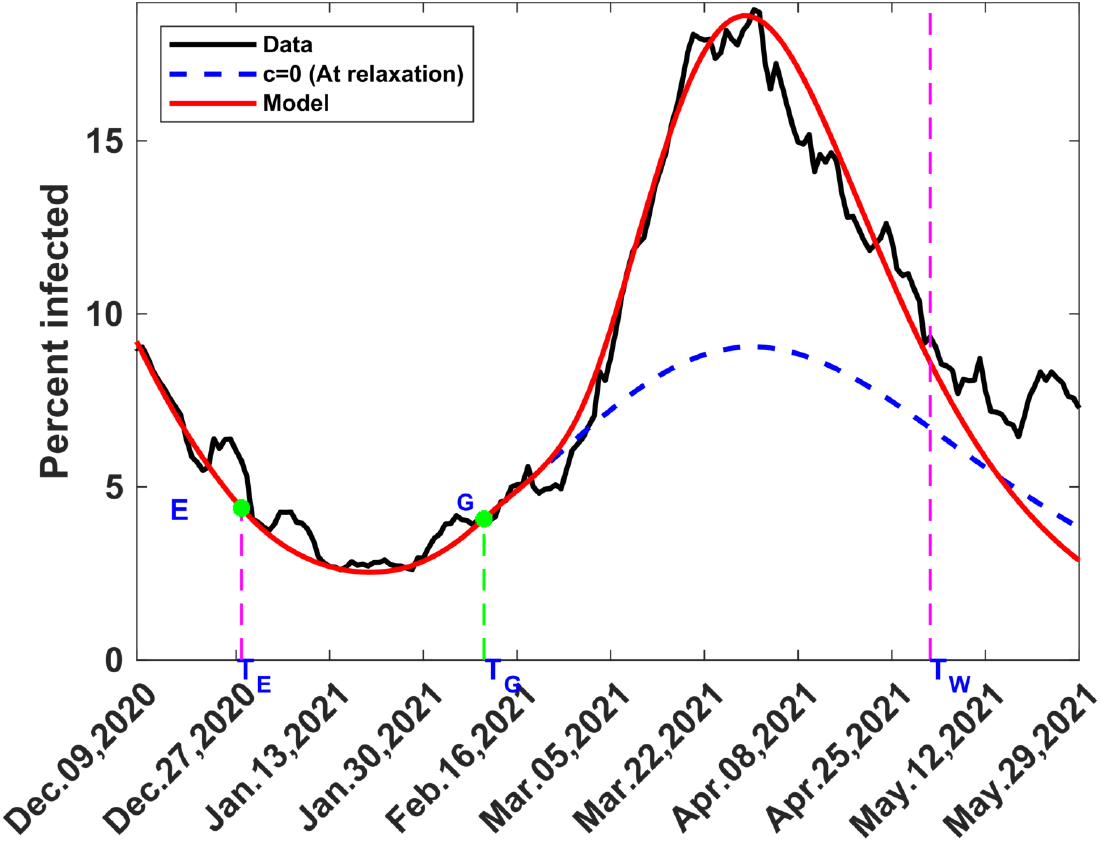
Computed and observed third wave of COVID-19 in Kenya, with the no-intervention curve shown in blue dashed curve. *E* represents the change point from second to the third wave and *T*_*E*_ the corresponding time; *G* is the point of application of delay function condition, at time *T*_*G*_; *T*_*W*_ represents the end of third wave.

#### 3.2.3 Fourth Wave

The 4^th^ wave of COVID-19 in Kenya appeared in two prominent spikes of different amplitudes. We modeled each spike separately before combining them to form the complete 4^th^ wave. For convenience of presentation, we will adopt some notations as follows. There is a change point from the 3^rd^ wave to the 1^st^ spike which we label *E*1; there is another change point from the 1^st^ to the 2^nd^ spike labeled as *E*2. We let *G*1 and *G*2 be the delay function points in the 1^st^ and 2^nd^ spikes, respectively, and *B*1 and *B*2 be the points for application of Equation (7) for values at *G*1 and *G*2, respectively.

**First Spike:** The methods in Section 2.4.2, led to *β*_*b*_ = 0.16339, *m* = 30, *c* = −7 (700% relaxation) and the 1^st^ change point, *T*_*E*1_ = 02-May-21. From Section 2.4.3, we identified *T*_*G*1_ = 01-Jun-21 and *I*_*G*1_ = 0.0724. Comparison with previous waves led to *T*_*B*1_ = 28-Feb-21 and *I*_*B*1_ = 0.0724. Equation (7) yielded the values in column 5, Section A2 of Table A1 The fore portion of the 1st spike is the arc *E*1*−G*1 . The methods in Section 2.4.4, applied at *T*_*V N*_ = *T*_*G*1_ = 01-Jun-21 with *β*_*b*_ = 1.3071, led to the non-intervention curve (blue dashed curve), predominantly above data as shown in Figure 7. Hence, we let *T*_*MT*_ = *T*_*V N*_ and undertook final mitigation for *t > T*_*MT*_ using *c* = 0.5 (50% mitigation) and *m* = 30. This gave the back portion of the 1st spike which, combined with the fore portion, yielded the complete 1^st^ spike in Figure 7.

**Figure 7:**
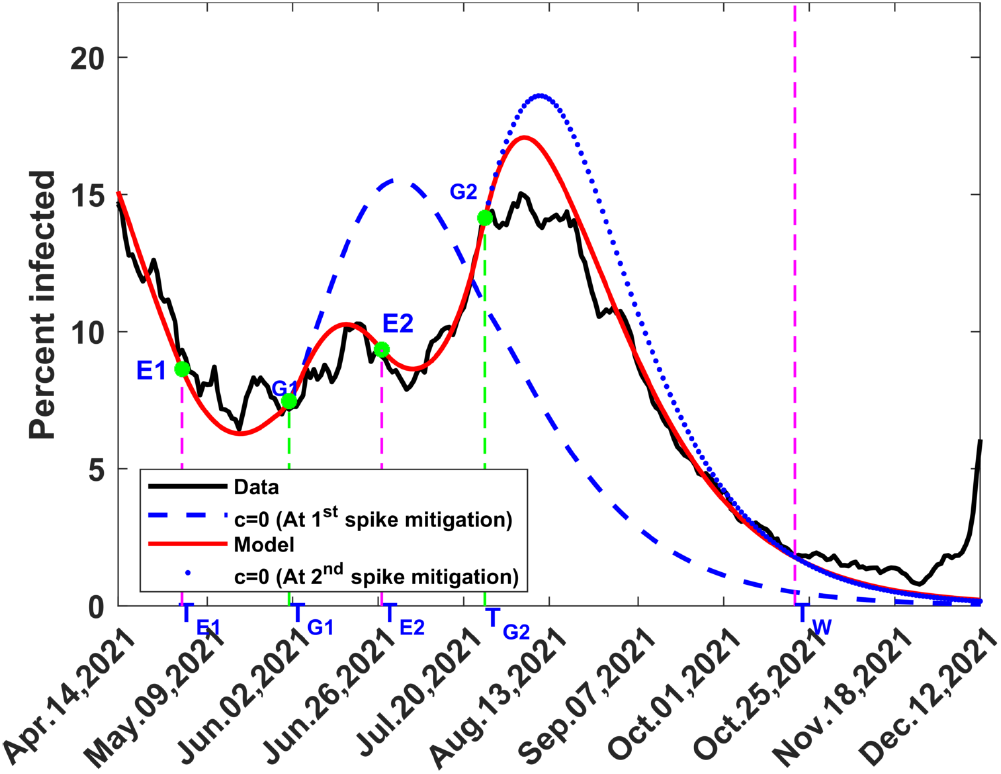
Computed and observed fourth wave of COVID-19 in Kenya. The blue dashed curve represents the no-intervention curve for the first spike while blue dotted curve represents the no-intervention curve for the second spike. *E*1 is the change point from 3^rd^ wave to the 1^st^ spike of the 4^th^ wave at time*T*_*E*1_; *G*1 is the point of application of delay function condition in the 1^st^ spike at time *T*_*G*1_, while *E*2 is the change point from 1^st^ to 2^nd^ spike at time *T*_*E*2_ and *G*2 the point of application of delay function condition in the 2^nd^ spike at time *T*_*G*2_; *T*_*W*_ is the end of the 4^th^ wave.

**Second Spike** The methods in Section 2.4.2, led to *β*_*b*_ = 0.079341, *m* = 30, *c* = −2.9 (290% relaxation). The 2^nd^ change point, *T*_*E*2_ = 27-Jun-21. From Section 2.4.3, we identified *T*_*G*2_ = 26-Jul-21 and *I*_*G*2_ = 0.143. Comparison with previous waves led to *T*_*B*2_ =14-Mar-21 and *I*_*B*2_ = 0.143. Equation (7) yielded the values in column 6, Section A2 of Table A1 The fore portion of the 2nd spike is the arc *E*2 *− G*2. The methods in Section 2.4.4, applied at *T*_*V N*_ = *T*_*G*2_ = 26-Jul-21 with *β*_*b*_ = 0.29571, led to the no-intervention curve blue dotted curve, predominantly above data as shown in Figure 7. Hence, we let *T*_*MT*_ = *T*_*V N*_ and undertook final mitigation for *t > T*_*MT*_ using *c* = 0.2 (20% mitigation) and *m* = 15. This gave the back portion of the 2nd spike which, combined with the fore portion, yielded the complete 2^nd^ spike as shown in Figure 7. Combination of the 1^st^ and 2^nd^ spikes led to the complete 4^th^ wave, in Figure 7.

#### 3.2.4 Fifth Wave

For convenience of presentation, we will adopt some notations as follows. There will be two delay function points which we label *G*1 and *G*2, respectively, and we let *B*1 and *B*2 be the points for application of Equation (7) for values at *G*1 and *G*2, respectively.

The methods in Section 2.4.2, led to *β*_*b*_ = 0.13071, *m* = 60, *c* = *−*10(1000% relaxation) and *TE* = 21-Oct-21. From Section 2.4.3, we identified *T*_*G*1_ = 02-Dec-21 and *I*_*G*1_ = 0.0147. Comparison with previous waves led to *T*_*B*1_ =27-Apr-20 and*I*_*B*1_ = 0.0147. Equation (7) yielded the values in column 7, Section A2 of Table A1. The methods in Section 2.4.4, applied at *T*_*V N*_ = *T*_*G*1_ = 02-Dec-21, led to the non-intervention curve (blue dashed curve), predominantly below data in Figure 8. Hence, we let *T*_*RX*_ = *T*_*V N*_ with *β*_*B*_ = 0.67286 and undertook relaxation for *t > T*_*RX*_ using *c* = −4(400% relaxation) and *m* = 15. The resulting model followed data for a while before tilting to the right. To realign the model with the data, we chose a 2nd delay function point, before the tilt, at *T*_*G*2=_14-Dec-21, with *I*_*G*2_ = 0.0988. Comparison with previous waves led to *T*_*B*2_ =16-Jul-21 and *I*_*B*2_ = 0.0985. Equation (7) yielded the values in column 7, Section A4 of Table A1. The fore portion of the wave is the arc *E − G*2. The methods in Section 2.4.3, applied at *T*_*V N*_ = *T*_*G*2_ =14-Dec-21 with *β*_*b*_ = 0.39473 led to the non-intervention curve (blue dotted curve), predominantly below data in Figure 8. Hence we let *T*_*RX*_ = *T*_*V N*_ = *T*_*G*2_ and undertook final relaxation for *t > T*_*RX*_ using *c* = −3.5(350% relaxation) and *m* = 5. This finalized the back portion of the wave, which, on combination with the fore portion, resulted in the complete 5th wave, in Figure 8.

**Figure 8:**
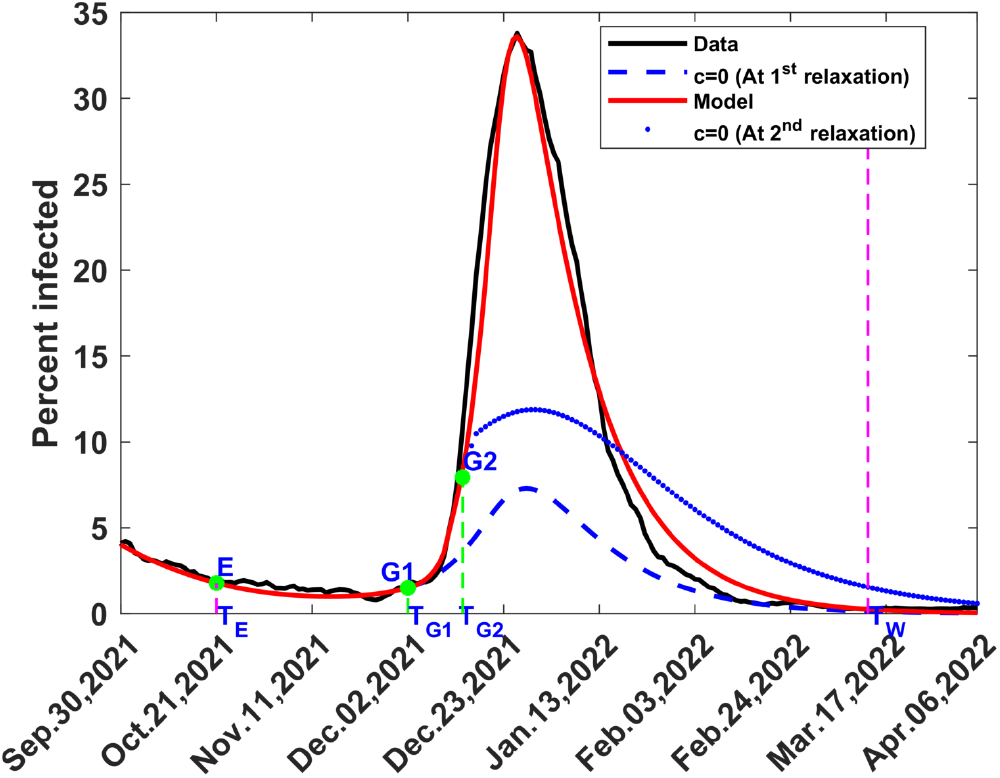
Computed and observed fifth wave of COVID-19 in Kenya. The dashed and dotted blue curves represent the non-intervention curves, red is the computed infections and black the data. E is the change point from 4^th^ wave to 5^th^ wave at time *T*_*E*_; G1 the first point of application of delay function condition at time *T*_*G*1_, and G2 the second point of application of delay function condition at time *T*_*G*2_; *T*_*W*_ is the end of 5^th^ wave.

### 3.3 Sixth and Seventh waves

The fore portions of the 6th and 7th waves were generated by exponential approximation, rather than by relaxation, as was the case with the previous waves. The results are, therefore, presented separately in this section.

#### 3.3.1 Sixth Wave

The methods in Section 2.5.1, led to *T*_*E*_ = 13-Mar-22, 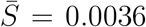, *β*_*XP*_ = 238 and *T*_*G*_ = 14-May-22. Equations (12) and (13) yielded disease variables from *T*_*E*_ to *T*_*G*_ such that *I*_*G*_ = 0.00817. Using Section 2.4.3, comparison with previous waves led to *T*_*B*_ =16-Apr-20 and *I*_*B*_ = 0.00827. Equation (7) yielded the values in column 3, Section A2 of Table A2. The fore portion of the wave is the arc *E − G*. The methods in Section 2.4.4, applied at *T*_*V N*_ = *T*_*G*_ =14-May-22, with *β*_*b*_ = 0.84538 led to the non-intervention curve (blue dashed curve), predominantly below data on the left side of Figure 9. Hence, we let *T*_*RX*_ = *T*_*V N*_ and undertook relaxation for *t > T*_*RX*_ using *c* = −0.15(15% relaxation) and *m* = 15. The solution yielded a model which closely fit the left hand side of the wave, as given in Figure 9. To complete the model, we effected another intervention at *T*_*V N*_ =17-Jun-22, near the apex of the wave. The methods in Section 2.4.4, applied at *T*_*V N*_ =17-Jun-22 with *β*_*b*_ = 0.169597 led to the non-intervention curve (blue dotted curve), predominantly above data on the right side of Figure 9. Hence we let *T*_*MT*_ = *T*_*V N*_ and undertook mitigation for *t > T*_*M*_ *T* using *c* = 0.82(82% mitigation) and *m* = 15. The solution completed the back portion of the curve which, on combination with the fore portion, resulted in the complete 6th wave, as given in Figure 9.

**Figure 9:**
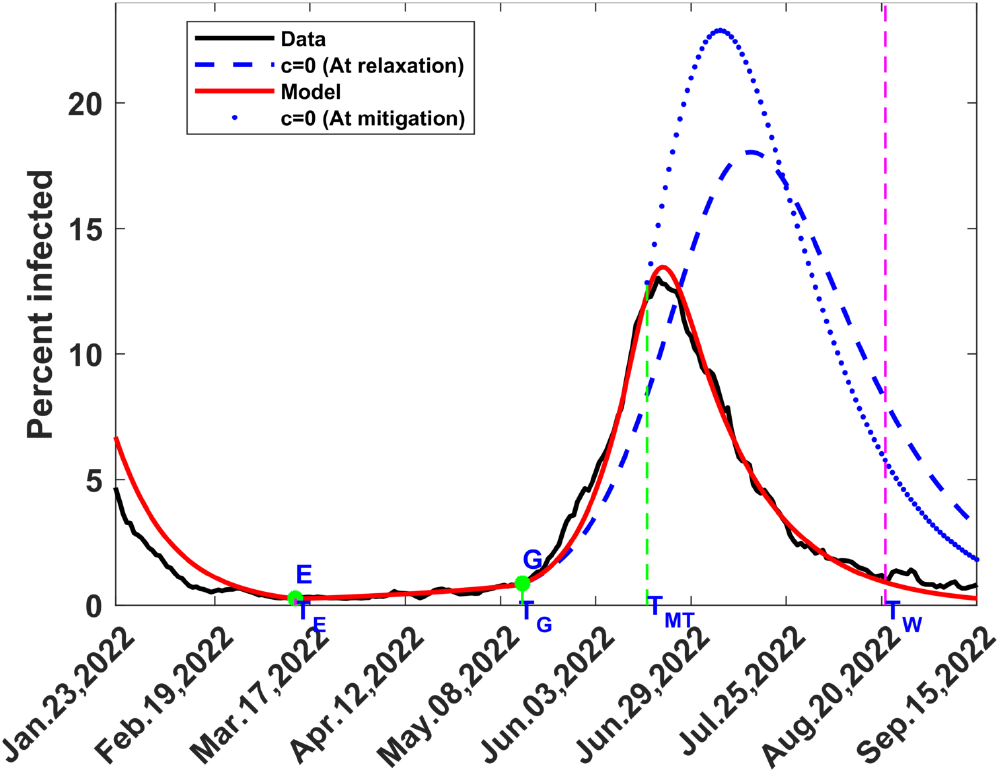
Computed and observed sixth wave of COVID-19 in Kenya. The blue curves represent the non-intervention curves, red the computed infections and black the data. *E* is the change point from 5^th^ to 6^th^ wave, with the corresponding time, *T*_*E*_, *G* the point of application of delay function condition at time *T*_*G*_, *T*_*MT*_ the time at which to apply mitigation and *T*_*W*_ the end of 6^th^ wave.

#### 3.3.2 Seventh Wave

The methods in Section 2.5.1, led to *T*_*E*_ = 22-Aug-22, 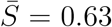, *β*_*XP*_ = 0.11 and *T*_*G*_ = 10-Oct-22. Equations (12)–(13) yielded disease variables from *T*_*E*_ to *T*_*G*_ such that *I*_*G*_ = 0.0101. Using Section 2.4.3, comparison with previous waves led to *T*_*B*_ =17-May-22 and *I*_*B*_ = 0.0105. Equation (7) yielded the values in column 4, Section A2 of Table A2. The fore portion of the wave is the arc *E −G*. The methods in Section 2.4.4, applied at *T*_*V N*_ = *T*_*G*_ =10-Oct-22, with *β*_*b*_ = 0.029875 led to the non-intervention curve (blue dashed curve), predominantly below data on the left side of Figure 10. Hence, we let *T*_*RX*_ = *T*_*V N*_ and undertook relaxation for *t > T*_*RX*_ using *c* = −0.2(20% relaxation) and *m* = 15. The solution yielded a model which closely fit the left hand side of the wave, as given in Figure 10. To complete the model, we effected another intervention at *T*_*V N*_ = 05-Nov-22, near the apex of the wave. The methods 2.4.4, applied at *T*_*V N*_ =05-Jun-22 with *β*_*b*_ = 0.1781 led to the non-intervention curve (blue dotted curve), predominantly above data on the right side of Figure 10. Hence we let *T*_*MT*_ = *T*_*V N*_ and undertook final mitigation for *t > T*_*MT*_ using *c* = 0.68(68% mitigation) and *m* = 10. The solution completed the back portion of the wave which, on combination with the fore portion, resulted in the complete 7th wave, as given in Figure 10. We noticed a lot of noise in the data from mid December 2022 before it stopped being posted in the public portal of the Ministry of Health website on 26 January 2023 [38].

**Figure 10:**
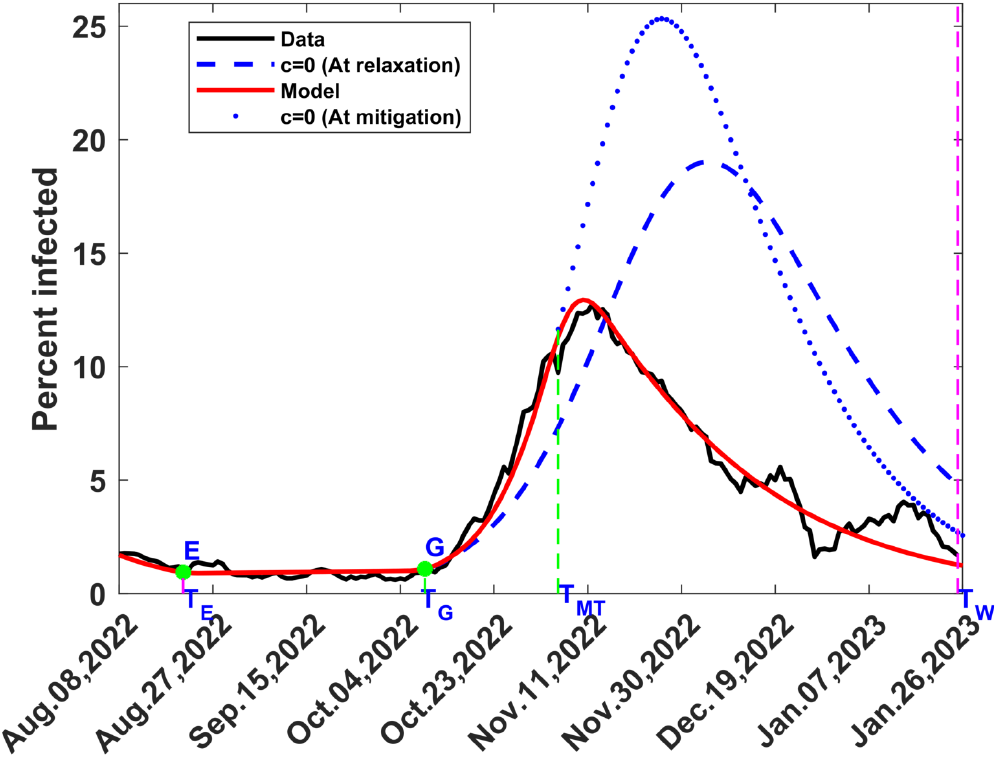
Computed and observed seventh wave of COVID-19 in Kenya. The blue dotted and dashed curves represent the non-intervention curves, red the computed infections and black the data. *E* is the change point from 6^th^ to 7^th^ wave at time *T*_*E*_, *G* the point of application of delay function condition at time *T*_*G*_, *T*_*MT*_ the time at which mitigation is applied and *T*_*W*_ the end of 7^th^ wave.

### 3.4 Complete COVID-19 waves in Kenya

Consolidation of Figure 2 and Figures 5 to 10, without the no-intervention curves, yields the complete COVID-19 waves in Kenya, as shown in Figure 11, where the model results are compared with data.

**Figure 11:**
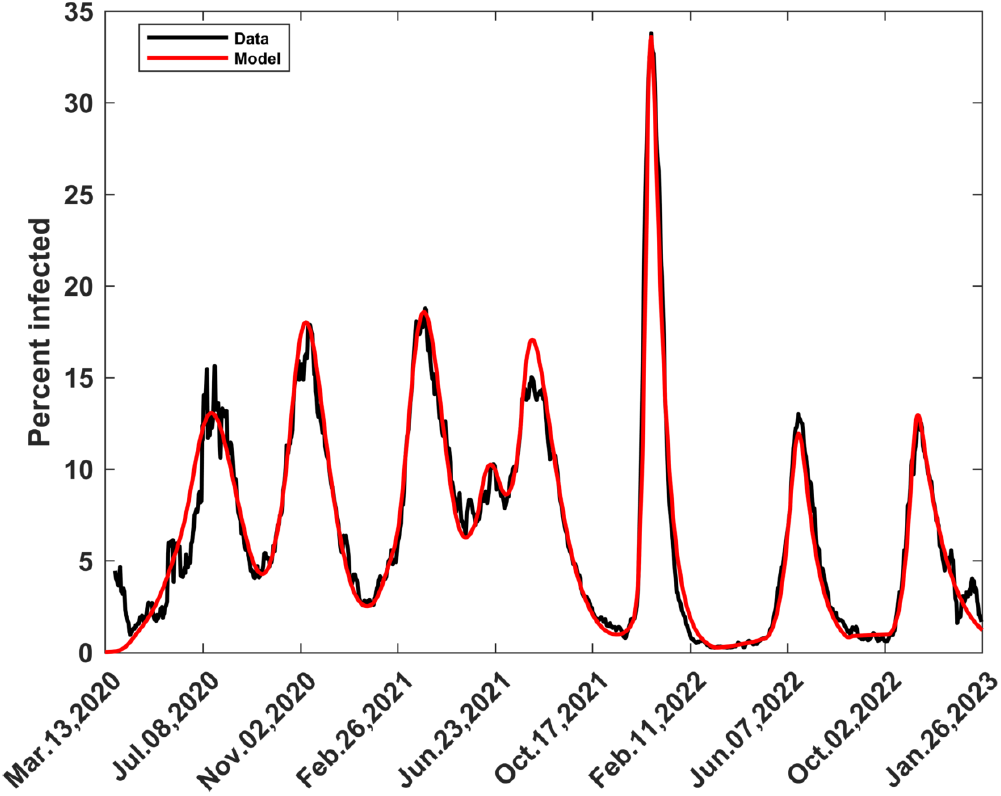
Computed and observed complete COVID-19 Waves in Kenya

In Table 1, we present the amplitudes and durations of the waves. The durations are based on the times between identified change points and may differ from those arrived at from clinical considerations [40]. The strongest wave was the 5th at 33.6% infected and the weakest was the 7th wave at 13.0% infected. The longest lasting was the 1st wave, with duration of 174 days while the shortest was the 2nd with duration of 116 days.

**Table 1:** Kenya COVID-19 wave amplitudes in 7-day averaged percent infected and durations in days.

|  | Wave 1 | Wave 2 | Wave 3 | Wave 4 | Wave 5 | Wave 6 | Wave 7 |
| --- | --- | --- | --- | --- | --- | --- | --- |
| Amplitude (% infected) | 13.1 | 18.0 | 18.6 | 17.1 | 33.6 | 13.6 | 13.0 |
| Duration, (Days) | 174 | 116 | 125 | 172 | 143 | 161 | 157 |

## 4 DISCUSSION AND CONCLUSIONS

In this article, we have formulated, analysed and computed a COVID-19 model that is based on the generation of two types of new waves. The first type is a wave generated from a vertically decreasing infection that diverges to the right, forms a bowl-like shape before increasing upwards, as shown in the second to the fifth wave; such a wave is modelled by exerting a sufficiently large relaxation force. Waves were generated by using relaxation ratios ranging from *c* = −2.9 (290% relaxation) to *c* = 10 (1000% relaxation). The stronger the wave the larger the relaxation ratio required, thus reflecting the force necessary to follow the contour of the wave sufficiently, before application of the delay function. The second type is a wave that emerges from very low infections that are nearly horizontal, as shown by the sixth and seventh waves; such a wave is modelled through an exponential infection, involving the transmission rate for exponential infection, *β*_*XP*_ ; this quantity had two vastly different values, namely 238 for the sixth wave and 0.11 for the seventh wave. There is no anomaly. The values are a result of choosing *β*_*XP*_ to satisfy Equation (11), with 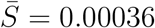 for the 6th wave and 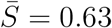 for the 7th wave. Although the models were the same within the two groups of waves, they were implemented to suit the characteristics of each wave. The results depicted in Figures 5 to 11 agree quite well with data, apart from areas where there are spikes, or there is noise, in the data. No other numerical results are available for comparison, for the complete COVID-19 waves in Kenya. Indeed the closest to this work are results for a few waves in India, by Gosh et. al. [32].

The change point that determines the transition from one wave to the next is obtained by noting the tail of the modelled preceding wave and where the trajectory of the new observed infection begins to diverge from this tail. This point of divergence forms an initial guess to the change point. The final change point is obtained after carrying out the procedures in Section 2.4.2 and will usually be within 2 weeks of the initial point. In Table A3, we have indicated the timeline of events relevant to COVID-19 dynamics in Kenya, including relaxation and mitigation events due to intervention. We have also indicated the months the variants of SARS-CoV-2 were dominant, as determined from genomic analysis [3, 40]. Finally, we have indicated the change points at which we decided to generate waves by application of large relaxation forces or through exponential approximation. The change points that we computed are within the months when genomic analyses showed particular variants of SARS-CoV-2 to be dominant; in most cases, they were actually at the beginning or in the middle of such months. This makes us conclude that the dominant variants of SARS-CoV-2 were the major sources of the relaxation forces that were capable of changing the infection trajectories. There were other relaxation forces relating to the lifting of, or noncompliance to, certain mitigation measures. In our view, such forces slightly enhanced the effects of the main drivers but they were not, on their own, sufficient to lead to generation of new waves.

The duration for optimum change in the transmission rate, m, had a value of 30 for waves whose bases took a shorter time to form, like the 2nd wave and spikes in the 4th wave. It had larger values for waves whose bases took longer to form, for instance 45 for the 3rd wave and 60 for the 5th. As the waves proceeded, their shapes were influenced by relaxation and mitigation forces from various sources, including interventions. This aspect was modelled through appropriate application of relaxation or mitigation by noting the position of the observed trajectory of infection relative to the no-intervention curve. The values of m here were either 10 or 15, apart from the 1st spike in the 4th wave for which it was 30. This implied that most interventions in the back portion of the wave resulted in the optimum change of the transmission rate being achieved close to the default value of 15 days.

We recommend extension of the developments in this article to investigations in several directions, as indicated hereunder:

1. Mathematical analyses to unravel the theory behind turning a decreasing contour of infection into an increasing contour through application of a large enough relaxation force. So far this observation is purely computational or numerical.
2. The method used to generate the 6th and 7th waves, combined with diligent monitoring, can be applied to detect a future wave of COVID-19 or other epidemic.
3. Application of smaller values of m, the optimum duration of optimum change of the transmission rate, together with use of no-intervention curve, could lead to detection of spikes of smaller amplitudes, although at a higher computational effort.
4. The methods described here can be used as a predictive tool if time series techniques are combined with computation of no-intervention curves.

## Data Availability

All data produced in the present study are available upon reasonable request to the authors

https://www.health.go.ke/

https://www.worldometers.info/coronavirus

## Acknowledgements

We acknowledge Alice Wangui Wachira, Anne Kanyua Kinyua and Lucy Nyanchama for their assistance with data collection.

## Data sources

All the data used is in the public domain [38],[39].

## Appendix A Wave computation and timeline of COVID-19 events

**Table A1:** Computing the 2nd to 5th COVID-19 Waves in Kenya.

| Variables and Parameters |  | 2nd Wave | 3rd Wave | 4th wave 1st Spike | 4th wave 2nd Spike | 5th Wave |
| --- | --- | --- | --- | --- | --- | --- |
| A. FRONT SECTION OF WAVES 2 - 5 |  |  |  |  |  |  |
| A1. Generation of new wave by relaxation |  |  |  |  |  |  |
| Change point | $T_E$ | 03-Sep-20 | 28-Dec-20 | 02-May-21 | 27-Jun-21 | 21-Oct-21 |
| Transmission rate preceding relaxation at $T_E$; $[= \beta(T_E)]$ | $\beta_b$ | 0.13507 | 0.17018 | 0.16339 | 0.079341 | 0.13071 |
| Relaxation ratio for wave generation | $c$ | -4 | -9 | -7 | -2.9 | -10 |
| Duration of optimal change of transmission rate | $m$ | 30 | 45 | 30 | 30 | 60 |
| Use values of $S, I, R, D$ and $\beta$ at $T_E$ to solve system till the time for enforcing delay-function as indicated below. | | | | | | |
| A2. Application of delay-function |  |  |  |  |  |  |
| Time in current wave at which delay-function is enforced | $T_G$ | 08-Oct-20 | 10-Feb-21 | 01-Jun-21 | 26-Jul-21 | 02-Dec-21 |
| Approximate infected ratio at $T_G$ | $I_G$ | 0.07394 | 0.04099 | 0.0724 | 0.143 | 0.0147 |
| Infection in previous wave that approximates $I_G$ | $I_B$ | 0.07394 | 0.04099 | 0.0724 | 0.143 | 0.0147 |
| Time at which $I_B$ occurs. | $T_B$ | 17-Jun-20 | 26-May-20 | 28-Feb-21 | 14-Mar-21 | 27-Apr-20 |
| Apply delay-function at $T_G$ to $S, I, R, D$ and $\beta$ | | | | | | |
| B. BACK SECTION OF WAVES 2 – 4 |  |  |  |  |  |  |
| [Intervention: Relaxation/Mitigation] |  |  |  |  |  |  |
| A.CONTINUATION OF FRONT SECTION FOR WAVE 5 |  |  |  |  |  |  |
| A3. Intervention (Relaxation) for Wave 5 |  |  |  |  |  |  |
| From $T_G$ solve the system for L days then apply relaxation | | | | | | |
| Number of days for solution after $T_G$ | L | 0 | 11 | 0 | 0 | 0 |
| Time at intervention ($= T_{RX}$ for $c < 0$ or $T_{MT}$ for $0 < c < 1$) | $T_{VN}$ | 08-Oct-20 | 21-Feb-21 | 01-Jun-21 | 26-Jul-21 | 02-Dec-21 |
| Transmission rate preceding intervention at $T_{VN}$; $[= \beta(T_{VN})]$ | $\beta_b$ | 0.67533 | 0.10892 | 1.3071 | 0.29571 | 0.67286 |
| Apply Relaxation/Mitigation from non-intervention curve | $c$ | -0.26 | -0.5 | 0.5 | 0.2 | -4 |
| Duration of optimal change of transmission rate | $m$ | 15 | 10 | 30 | 15 | 15 |
| For waves 1 to 4, use the values at $T_{VN}$ to solve the system for $t>T_{-}\{VN\}$ and obtain the rest of the wave. For 5th wave the process continues as indicated below. | | | | | | |
| A4. Application of Second delay-function for Wave 5 |  |  |  |  |  |  |
| L days from relaxation, apply another delay-function |  |  |  |  |  |  |
| Number of days to follow previous relaxation | L | — | — | — | — | 12 |
| Time at which the 2nd delay-function is enforced | $T_{G2}$ | — | — | — | — | 14-Dec-21 |
| Approximate infected fraction at $T_{G2}$ | $I_{G2}$ | — | — | — | — | 0.0988 |
| Infection in previous wave that approximates $I_{G2}$ | $I_{B2}$ | — | — | — | — | 0.0985 |
| Time in previous wave where $I_{B2}$ occurs. | $T_{B2}$ | — | — | — | — | 16-Jul-21 |
| Apply delay-function at $T_{B2}$; Follow by relaxation at $T_{-}\{B2\}$. | | | | | | |
| C. BACK SECTION OF WAVE 5 |  |  |  |  |  |  |
| [Intervention: Relaxation] |  |  |  |  |  |  |
| Time for 2nd relaxation ($= T_{G2}$) | $T_{RX}$ | — | — | — | — | 14-Dec-21 |
| Transmission rate preceding 2nd relaxation; $[= \beta(T_{RX})]$ | $\beta_b$ | — | — | — | — | 0.39473 |
| Relaxation ratio | $c$ | — | — | — | — | -3.5 |
| Duration of optimal change of transmission rate | $m$ | — | — | — | — | 5 |
| Solve the system for $t > T_{RX}$ and obtain the rest of the wave. | | | | | | |

**Table A2:**
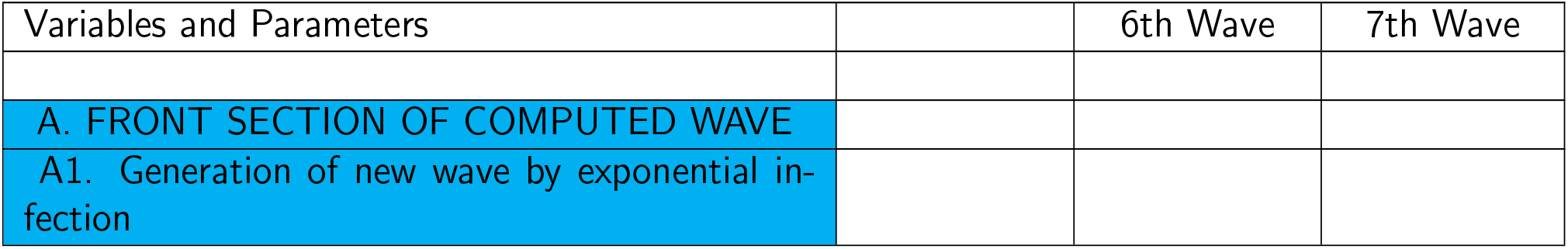
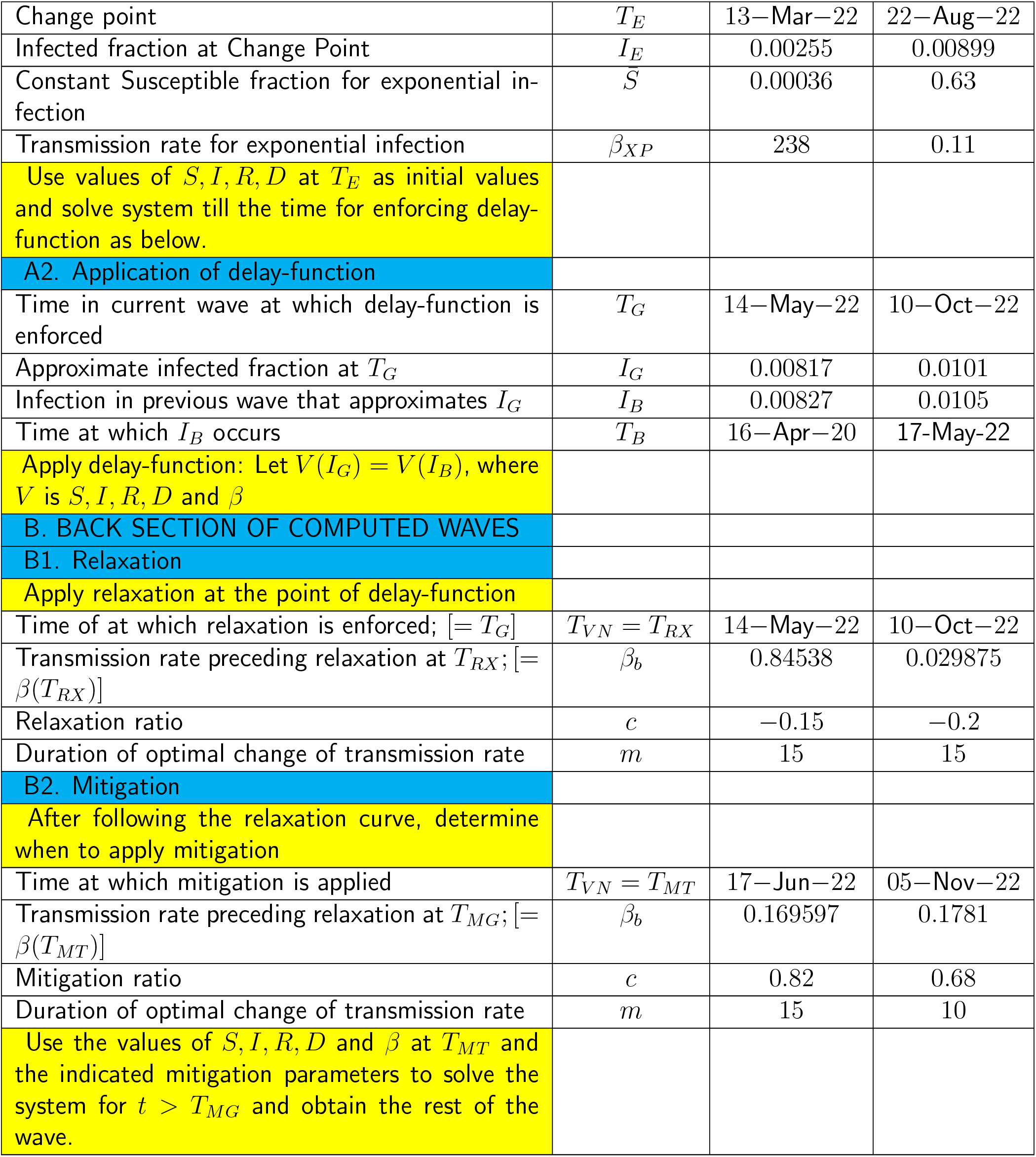
Computing the 6th and 7th COVID-19 Waves in Kenya.

**Table A3:** Timeline of COVID-19 events in Kenya.

|  |  |
| --- | --- |
| WAVE: 1st | DATES: MAR-2020 to SEP-2020 \| CHANGE POINT: 13-Mar-20 |
| MITIGATION: | Closure of learning institutions; Restrictions on movement, restaurants, public transport, political, religious and social gatherings; Nation-wide curfew; Lockdown in some cities; Public hygiene & social distancing; Air travel suspended. |
| RELAXATION: | 1st COVID-19 wave; Dominant SARS-CoV-2 variant - B.1.<br>Lockdown & restriction on movement lifted; Air travel resumed; Other controls relaxed. |
| WAVE: 2nd | DATES: SEP-2020 to NOV-2020 \| CHANGE POINT: 03-Sep-20 |
| MITIGATION: | Restrictions imposed again on restaurants, clubs, social and religious gatherings. |
| RELAXATION: | 2nd COVID-19 wave; Dominant SARS-CoV-2 variant – Beta;<br>Restrictions eased on curfew, restaurants, social & religious gatherings; Schools partially open. |
| WAVE: 3rd | DATES: DEC-2020 to APR-2021 \| CHANGE POINT: 28-Dec-20 |
| MITIGATION: | Restrictions on restaurants, social & religious gatherings, public transport;<br>Lockdown of 5 counties (Nairobi and its environs); Vaccination starts – target of 27.25 million. |
| RELAXATION: | 3rd COVID-19 wave; Dominant SARS-CoV-2 variant - Alpha;<br>Opening of learning institutions; Relaxation in public transport protocols. |
| WAVE: 4th | DATES: MAY-2021 to OCT-2021 \| CHANGE POINT: 02-May-21 |
| MITIGATION: | Country-wide curfew; Restrictions on social gatherings; Vaccination (19.5% of target) |
| RELAXATION: | 4th COVID-19 wave; Dominant SARS-CoV-2 variant - Delta;<br>Lockdown lifted of 5 counties (Nairobi and its environs). |
| WAVE: 5th | DATES: OCT-2021 to FEB-2022 \| CHANGE POINT: 21-Oct-21 |
| MITIGATION: | Enhanced vaccination campaign (45.7% of target). |
| RELAXATION: | 5th COVID-19 wave; Dominant SARS-CoV-2 variant - Omicron;<br>Country-wide curfew lifted; Holiday travels; Learning institutions open. |
| WAVE: 6th | DATES: MAR-2022 to AUG-2022 \| CHANGE POINT: 13-Mar-22 |
| MITIGATION: | Enhanced vaccination campaign (64.8% of target) |
| RELAXATION: | 6th COVID-19 wave; Dominant SARS-CoV-2 variant – Omicron, sub-variants BA.4 & BA.5;<br>Election campaigns; Safari rally. |
| WAVE: 7th | DATES: AUG-2022 to JAN-2023 \| CHANGE POINT: 22-Aug-22 |
| MITIGATION: | Enhanced vaccination campaign (84.7% of target). |
| RELAXATION: | 7th COVID-19 wave; Dominant SARS-CoV-2 variant - Omicron BQ1 and BQ1.1 ;<br>Election campaigns; Holiday travels; Learning institutions open. |

